# Role of the common *PRSS1-PRSS2* haplotype in alcoholic and non-alcoholic chronic pancreatitis: meta- and re-analyses

**DOI:** 10.1101/2020.10.13.20212365

**Authors:** Anthony F. Herzig, Emmanuelle Génin, David N. Cooper, Emmanuelle Masson, Claude Férec, Jian-Min Chen

## Abstract

The association between a common *PRSS1-PRSS2* haplotype and alcoholic chronic pancreatitis (ACP), which was revealed by the first genome-wide association study of chronic pancreatitis (CP), has been consistently replicated. However, the association with non-ACP (NACP) has been controversial. Herein, we sought to clarify this basic issue by means of an allele-based meta-analysis of currently available studies. We then used studies informative for genotype distribution to explore the biological mechanisms underlying the association data and to test for gene-environment interaction between the risk haplotype and alcohol consumption by means of a re-analysis. A literature search was conducted to identify eligible studies. Meta-analysis was performed using the Review Manager software. The association between the risk genotypes and NACP or ACP was tested for the best-fitting genetic model. Gene-environment interaction was estimated by both case-only and multinomial approaches. Five and eight studies were employed for the meta-analysis of ACP and NACP findings, respectively. The risk allele was significantly associated with both ACP (pooled OR 1.67, 95% CI 1.56–1.78; *P*<0.00001) and NACP (pooled OR 1.28, 95% CI 1.17–1.40; *P*<0.00001). Consistent with a dosage effect of the risk allele on *PRSS1*/*PRSS2* mRNA expression in human pancreatic tissue, both ACP and NACP association data were best explained by an additive genetic model. Finally, the risk haplotype was found to interact synergistically with ACP.

## INTRODUCTION

Chronic pancreatitis (CP) is a chronic inflammatory process of the pancreas that leads to irreversible morphological changes and progressive impairment of both exocrine and endocrine functions (1, 2). CP can be caused by genetic and/or environmental factors (3). In the one extreme, rare gain-of-function missense or copy number variants in the *PRSS1* gene (encoding cationic trypsinogen; MIM# 276000) can cause autosomal dominant hereditary pancreatitis (4, 5). In the other extreme, alcohol abuse is the most frequent environmental factor causing CP worldwide (2, 3).

Eight years ago, the first genome-wide association study of CP was published; a common *PRSS1-PRSS2* haplotype was reported to be associated with both alcoholic CP (ACP) and non-ACP (NACP) (6). *PRSS2* encodes anionic trypsinogen (MIM# 601564), the second major trypsinogen isoform after cationic trypsinogen. This association has a strong biological basis: the risk [C] allele of the lead single nucleotide polymorphism (SNP), rs10273639C/T, appeared to be associated with increased *PRSS1* mRNA expression in pancreatic tissue in a dosage-dependent manner (6). rs10273639C/T is in perfect linkage disequilibrium with rs4726576C/A, which is located 204 bp upstream of the translation initiation codon of the *PRSS1* gene; the C allele of rs4726576C/A increased reporter gene expression as determined by an *in vitro* promoter assay (7). Uncontrolled trypsin activity is central to CP pathogenesis (8).

Another interesting finding from the Whitcomb study was that rs10273639C/T was associated with a stronger genetic effect in ACP than in NACP, suggesting that the SNP-conferred risk was amplified by alcohol consumption. Moreover, in a case-only analysis, the SNP seemed to interact with ACP (6). In subsequent studies, the association of the common *PRSS1-PRSS2* haplotype with ACP has been consistently replicated. However, the association with NACP was equivocal, raising the possibility that the common *PRSS1-PRSS2* haplotype modifies CP risk only for ACP but not in NACP (9, 10). To explore this basic question and the underlying biological mechanisms, we firstly performed an allele-based meta-analysis of the reported associations between the common *PRSS1-PRSS2* haplotype and ACP and NACP. We then used informative studies in terms of genotype distribution to perform a new re-analysis with respect to (i) the fit of different genetic models to the association data and (ii) the gene-environment interaction between the common *PRSS1-PRSS2* haplotype and alcohol consumption status in CP.

## MATERIALS AND METHODS

### Literature search and inclusion criteria

The literature search was performed by keyword search (*PRSS1* and Pancreatitis) via PubMed (as of 13 July 2020), which was complemented by perusal of references citing the Whitcomb and/or Derikx studies (6, 9) via Google Scholar. Only studies that satisfied the following criteria were included for meta-analysis: (i) published in a peer-reviewed journal; (ii) the common *PRSS1-PRSS2* haplotype was analyzed in both ACP and/or NACP patients and controls; and (iii) the tagging SNP of interest was in Hardy–Weinberg equilibrium in the controls.

### Data extraction

The protective alleles of the SNPs that tagged the common *PRSS1-PRSS2* haplotype have often been used for statistical analysis in previous publications. To consider the genetic effects of both rare and common *PRSS1* variants in the same framework, herein we used the risk alleles of the tagging SNPs for analysis. To this end, risk allele or genotype distribution data in patients and controls were manually extracted from included studies.

### Statistical analysis and meta-analysis with respect to risk allele frequencies in patients and controls

The assessment of significance of the differences between the frequencies of the risk-tagging SNPs in patients and controls in the context of each study was performed by means of χ^2^ tests in R (11). A difference was regarded as being statistically significant when the *P* value was ≤0.05. The sample size for case-control association studies was estimated using the Online Sample Size Estimator (12).

Meta-analysis, heterogeneity and sensitivity analyses, and forest plots were all performed using the Review Manager 5.3 software (13). Funnel plots were not performed due to the small number of eligible studies (13, 14). The Mantel-Haenszel fixed effect or random effect model was used to compute pooled odds ratio (OR) in the absence or presence of statistical heterogeneity. Heterogeneity was considered to be significant when the *P* value for the test of heterogeneity was <0.05 or I^2^ was 50% or more (13, 15). The conduct and reporting of the meta-analysis were essentially in accordance with the PRISMA guidelines (16). This meta-analysis was not registered in PROSPERO (17), which does not allow the registration of already finished studies.

### Evaluation of the effect of rs10273639 on the expression of *PRSS1* and *PRSS2* in pancreatic tissue

This analysis was performed using the Genotype-Tissue Expression (GTEx) dataset (18).

### Re-analysis for association between the common *PRSS1-PRSS2* haplotype and ACP/NACP under different genetic models

Logistic regression models were fitted to examine the association between the risk genotypes and NACP or ACP in the context of single population data (refer to Results). Four models were tested: the genotypes were coded as either recessive, dominant, additive, or as each having an independent effect (general model) (19). We evaluated the Akaike information criterion (AIC) for each model and tested the change in deviance of the recessive, dominant, and additive models against the general model. Pearson’s residuals were calculated to ensure the reasonability of the models fitted. All analyses were carried out using the statistical analysis software R (version 3.6.1) (11).

### Re-analysis to test for interaction between the common *PRSS1-PRSS2* haplotype and alcohol consumption status in CP

The studies included in this analysis were those for which we could ascertain individual level genotype data for ACP, NACP and healthy control individuals. We firstly performed case-only analyses before combining results through meta-analysis in order to estimate a pooled OR for the interaction term (also referred to as the synergy index SIM). This analysis was performed using the metagen function from the meta R package (20).

In a second step, we applied the multinomial approach to test for gene-environment interactions as proposed by Kazma and colleagues (21). ACP and NACP patients were compared using the control individuals as a common baseline through a multinomial logistic regression model (multinom function from the nnet R package) (22). This method of investigating the gene-environment interaction is equivalent to the case-only approach (23); testing the interaction on a multiplicative scale (24). However, unlike the case-only approach, the multinomial model allows us to also include a control group and hence to simultaneously estimate the genetic effects and test for the presence of an interaction term, thereby giving a more complete description.

## RESULTS

### Included studies for meta-analysis

The keyword search (*PRSS1* and Pancreatitis) via PubMed resulted in 173 entries starting from the pioneering Whitcomb study (6). Of these, 15 publications were identified to report original data on the frequency of the common *PRSS1-PRSS2* haplotype in both pancreatitis patients and controls. Two studies that analyzed patients with a known non-alcohol-related external factor (25, 26) and three studies that analyzed only patients with acute pancreatitis (27-29) were immediately excluded from further consideration. Two additional reports that analyzed chronic pancreatitis (CP) patients were also excluded: one (30) because of a significant overlap of patients with a previous study (9) and the other due to its small sample size (i.e., only 85 CP cases and 78 controls were successfully genotyped) and lack of information on the numbers of CP subtypes among the analyzed patients (31). The remaining eight studies were included in our meta-analysis; five of these reported data on both alcoholic CP (ACP) and non-ACP (NACP) (6, 9, 10, 32, 33) whereas the other three reported data only for NACP (34-36). The search and selection processes of the included studies are outlined in Figure 1. In all included studies, the tagging single nucleotide polymorphism (SNP) of interest was either described in the original publication or verified here to be in Hardy–Weinberg equilibrium in the controls. Additionally, all included studies were evaluated to have a Newcastle-Ottawa Scale (NOS) score (37) of ≥7. Finally, perusal of references citing the Whitcomb study (6) and/or the Derikx study (9) via Google Scholar did not identify additional eligible studies in peer-reviewed journals.

**Figure 1.**
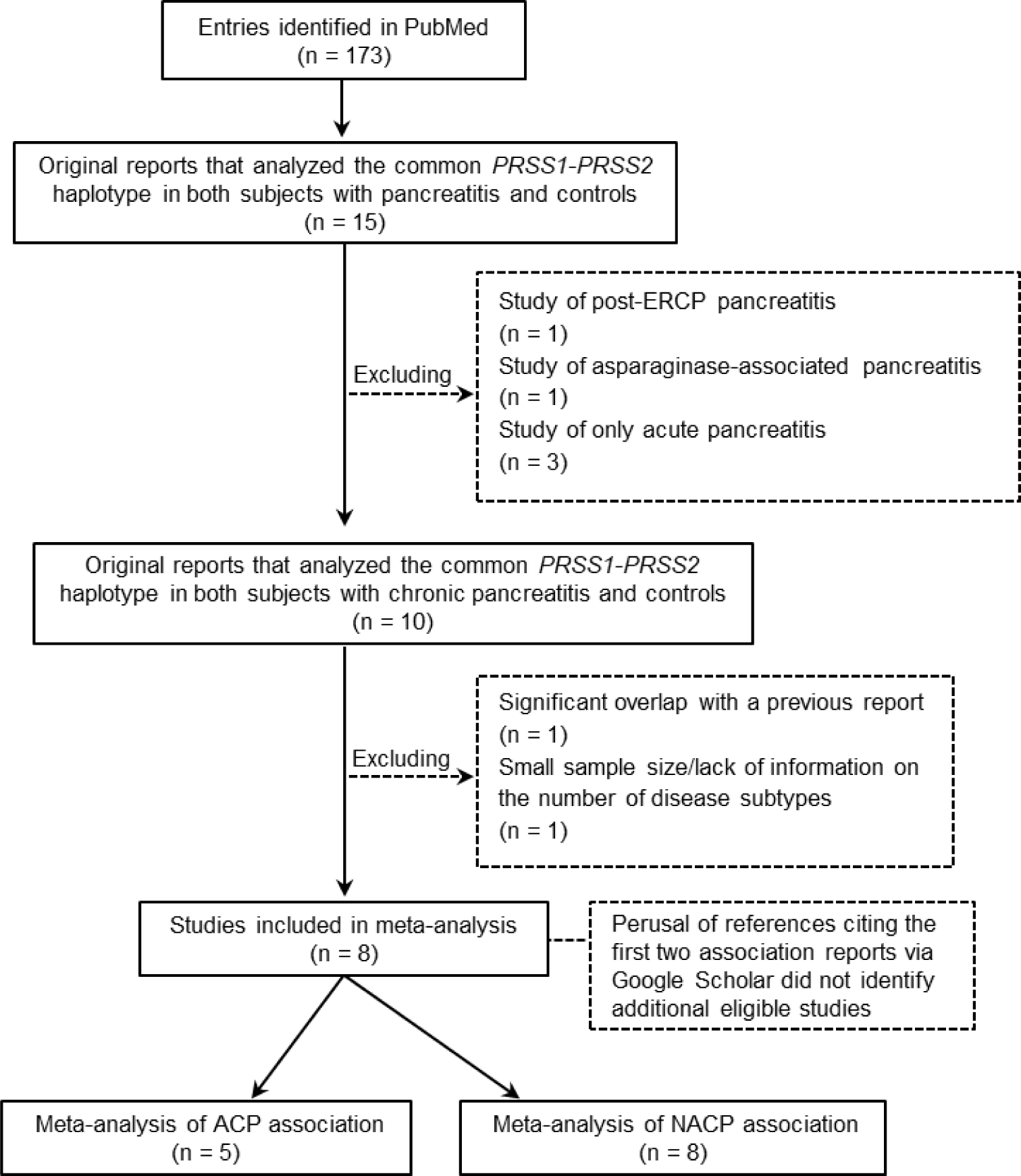
Flow chart of the search and selection process for the studies used for the allele-based meta-analysis. ACP, alcoholic chronic pancreatitis. ERCP, endoscopic retrograde cholangio-pancreatography. NACP, non-alcoholic chronic pancreatitis.

Basic characteristics of the eight included studies are summarized in Table 1. It should be noted that, in the Whitcomb study, the ACP and NACP cohorts actually comprised some patients with recurrent acute pancreatitis; CP and recurrent acute pancreatitis were modeled together based upon the assumption that the two disease states have common susceptibilities (6). As opined by the original authors, this treatment may reduce power relative to that of the analysis of CP alone but is unlikely to have altered the main conclusions of the study.

**Table 1.** Basic characteristics of the included studies and corresponding risk allele frequencies for the common *PRSS1*-*PRSS2* haplotype-tagging SNPs in patients and controls

| Study | Study population(s) | Risk allele of the tagging SNP | Number of patients/controls | Allele frequency in patients | Allele frequency in controls | OR (95% CI) | P value ($\chi^2$ test) |
| --- | --- | --- | --- | --- | --- | --- | --- |
| **ACP** |  |  |  |  |  |  |  |
| Whitcomb et al. (2012)<sup>6</sup> | Mainly Americans with European ancestry and a small number of German and British subjects | rs10273639[C] | 447<sup>a</sup>/8029 | 69.6% (622/894) | 57.6% (9249/16058) | 1.68 (1.45 – 1.95) | 2.03e-12 |
| Derikx et al. (2015)<sup>9</sup> | European (from 9 countries) | rs10273639[C] | 1854/5065 | 69.0% (2560/3708) | 57.6% (5835/10130) | 1.64 (1.52 – 1.78) | 2.20e-16 |
| Masamune et al. (2015)<sup>32</sup> | Japanese | rs10273639[C] | 272/480 | 32.5% (177/544) | 22.2% (213/960) | 1.69 (1.34 – 2.14) | 1.43e-5 |
| Giri et al. (2016)<sup>33</sup> | Indian | rs2855983[G] | 85/1288 | 48.8% (83/170) | 34.0% (876/2576) | 1.85 (1.36 – 2.53) | 0.00012 |
| Hegyi et al. (2020)<sup>10</sup> | Hungarian | rs6666[C] | 120/296 | 70.0% (168/240) | 57.3% (339/592) | 1.74 (1.26 – 2.40) | 0.00086 |
| **NACP** |  |  |  |  |  |  |  |
| Whitcomb et al. (2012)<sup>6</sup> | Mainly Americans with European ancestry and a small number of German and British subjects | rs10273639[C] | 1129<sup>b</sup>/8029 | 63.4% (1431/2258) | 57.6% (9249/16058) | 1.27 (1.16 – 1.40) | 2.10e-7 |
| Derikx et al. (2015)<sup>9</sup> | European (from three countries) | rs10273639[C] | 1192/4323 | 59.6% (1421/2384) | 57.6% (4981/8646) | 1.09 (0.99 – 1.19) | 0.085 |
| Masamune et al. (2015)<sup>32</sup> | Japanese | rs10273639[C] | 197/480 | 30.5% (120/394) | 22.2% (213/960) | 1.54 (1.18 – 2.00) | 0.0017 |
| Avanthi et al. (2015)<sup>34</sup> | Indian | rs10273639[C] | 96/156 | 30.7% (59/192) | 28.9% (90/312) | 1.09 (0.74 – 1.62) | 0.73 |
| Giri et al. (2016)<sup>33</sup> | Indian | rs2855983[G] | 434/1288 | 42.0% (364/868) | 34.0% (876/2576) | 1.40 (1.20 – 1.64) | 3.07e-5 |
| Paliwal et al. (2016)<sup>35</sup> | Indian | rs10273639[C] | 551/801 | 33.8% (372/1102) | 26.9% (431/1602) | 1.38 (1.17 – 1.64) | 0.00015 |
| Campa et al. (2018)<sup>36</sup> | European (from 8 countries) | rs10273639[C] | 345/4580 | 64.4% (444/690) | 57.8% (5292/9160) | 1.32 (1.12 – 1.55) | 0.00085 |
| Hegyi et al. (2020)<sup>10</sup> | Hungarian | rs6666[C] | 103/296 | 63.1% (130/206) | 57.3% (339/592) | 1.28 (0.92 – 1.77) | 0.17 |

<sup>a</sup>Including 113 patients with recurrent acute pancreatitis.<sup>b</sup>Including 462 patients with recurrent acute pancreatitis.
Abbreviations: ACP, alcoholic chronic pancreatitis; CI, confidence interval; NACP, non-alcoholic chronic pancreatitis; OR, odds ratio; SNP, single nucleotide polymorphism.

### Allele-based meta-analysis confirmed an association of the common *PRSS1-PRSS2* haplotype with both ACP and NACP

Although the frequencies of the risk-tagging alleles differ significantly (from 22% to 58%) between the American (primarily of European ancestry), European, Japanese, Indian and Hungarian control populations studied, all five ACP studies found a statistically significant association between the common *PRSS1-PRSS2* haplotype and ACP with remarkably comparable ORs (i.e., 1.64 to 1.85; Table 1). Application of the test for heterogeneity confirmed that the five studies were homogeneous (χ^2^ = 0.68, *P* = 0.95; I^2^ = 0%). The pooled OR (under a fixed effect model) was 1.67 (95% CI 1.56–1.78; *P* < 0.00001) (Figure 2).

**Figure 2.**
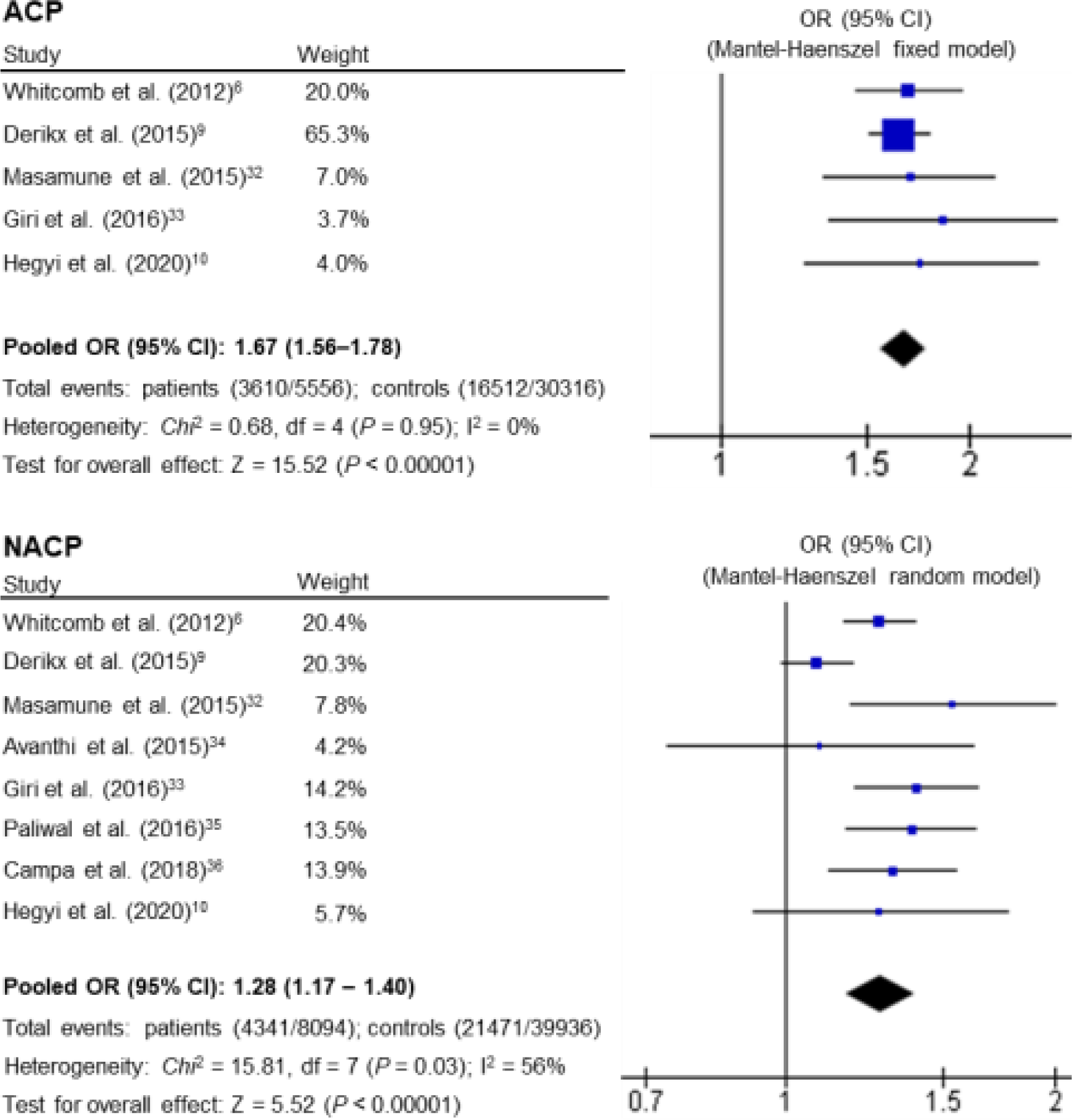
Results of the allele-based meta-analysis of the association between the common *PRSS1-PRSS2* haplotype and ACP or NACP, and the corresponding forest plots. Total events in patients and controls: number of risk alleles/number of total alleles. See Table 1 for the risk allele distribution data, OR (95% CI) and *P* values in the context of each individual study. ACP, alcoholic chronic pancreatitis. CI, confidence interval. NACP, non-alcoholic chronic pancreatitis. OR, odds ratio.

Of the eight NACP studies, five found a statistically significant association with NACP whereas the other three did not (Table 1). The test for heterogeneity revealed significant heterogeneity across the studies (χ^2^ = 15.81, *P* = 0.03; I^2^ = 56%). Sensitivity analysis identified the large European study (9) to be the source of the heterogeneity. Using a random-effect model, the common *PRSS1-PRSS2* haplotype showed significant association with NACP: the pooled OR was 1.28 (95% CI 1.17–1.40; *P* < 0.00001) (Figure 2).

The three studies that failed to show a significant association with NACP nevertheless all yielded ORs >1 (Table 1). Notably, the frequency of the tagging rs6666[C] allele among the Hungarian NACP patients was nearly 6% higher than in controls (i.e., 63.1% vs. 57.3%), raising the possibility that the absence of an association (*P* = 0.17) in the corresponding study (10) might have been due to the relatively small sample size (only 103 patients and 296 controls were examined). Assuming (i) the frequencies of the rs6666[C] allele in the Hungarian NACP patients and controls were 63% and 57%, respectively and (ii) a 1:2 ratio of cases to controls, at least 784 patients and 1568 controls would have been required to achieve significance at the 5% level with 80% power.

As for the large European study (9), the risk allele frequency in the NACP patients was 2% higher than in the controls, yielding a *P* value of 0.085 (Table 1). This large study used three NACP cohorts, viz. German, French and Dutch. The German cohort was the largest and showed a statistically significant association with NACP (OR = 1.21, p = 0.0027; Table 2).

**Table 2.** Comparison of the rs10273639[C] allele frequencies in the NACP patients and controls in the context of each population studied by Derikx et al. (ref. 9)

| Population | Number of patients/controls | All frequency in patients | Allele frequency in controls | OR (95% CI) | *P* value ($\chi^2$ test) |
| --- | --- | --- | --- | --- | --- |
| German | 690/2825 | 62.97% (869/1380) | 58.50% (3305/5650) | 1.21 (1.07 – 1.36) | 0.0027 |
| French | 415/1064 | 54.10% (449/830) | 54.61% (1162/2128) | 0.98 (0.83 – 1.15) | 0.83 |
| Dutch | 87/434 | 59.20% (103/174) | 59.22% (514/868) | 1.00 (0.72 – 1.39) | 1.00 |

Abbreviations: OR, odds ratio; CI, confidence interval; NACP, non-alcoholic chronic pancreatitis.

The third study that failed to achieve a significant association (34) was the smallest of the three Indian studies (Table 1).

### The risk allele number is positively correlated with the pancreatic *PRSS2* mRNA expression level

We evaluated whether the risk [C] allele of rs10273639C/T is associated with increased trypsinogen gene expression in the pancreas in the GTEx dataset. No significant expression quantitative trait loci (eQTLs) for *PRSS1* in the pancreas are currently available in GTEx. However, the genotypes of rs10273639 affected *PRSS2* mRNA expression in the pancreas in a dosage-dependent manner (see Supplementary Figure S1).

### The additive genetic model best fits the associations of the common *PRSS1-PRSS2* haplotype with NACP and ACP

The positive correlation between the risk allele number of rs10273639 and *in vivo PRSS1* and *PRSS2* mRNA expression implies that an additive genetic model would best explain the association data. To test this postulate, we firstly examined the three possible genetic models, namely dominant, recessive and additive, in the context of three NACP studies informative for genotype distribution, each from a single population (i.e., the only Japanese study (32), the Paliwal study that analyzed the largest number of Indian patients (35), and the German cohort (the largest of the European cohorts) studied by Derikx et al. (9)). A general model with an independent effect of each genotype was also fitted to aid the evaluation of the three models of interest.

The distribution of the rs10273639 genotypes showed significant differences between patients and controls under all three genetic models (Table 3). Pearson’s residuals (which should not greatly exceed a value of 2) were calculated to establish that all models had provided a reasonable description of the data. By evaluating the Akaike information criterion (AIC), we observed that the best fitting model was invariably the additive model as a lower AIC represents a better model fit. On examining the general models, we observed that the coefficients for the CC genotype were very close to twice the coefficients of the CT genotype (i.e., 0.39 vs. 0.25 in the German data; 0.87 vs. 0.45 in the Japanese data; and 0.59 vs. 0.34 in the Indian data). This is indicative of an additive dose-dependence relationship between the genotypes and the risk of CP. Further, we performed analyses of variance (Anova) of the recessive, dominant and additive models against the general model. This showed that only the recessive model provided a worse description of the data compared to the general model (*P* values below 0.05) (Table 3).

**Table 3.**
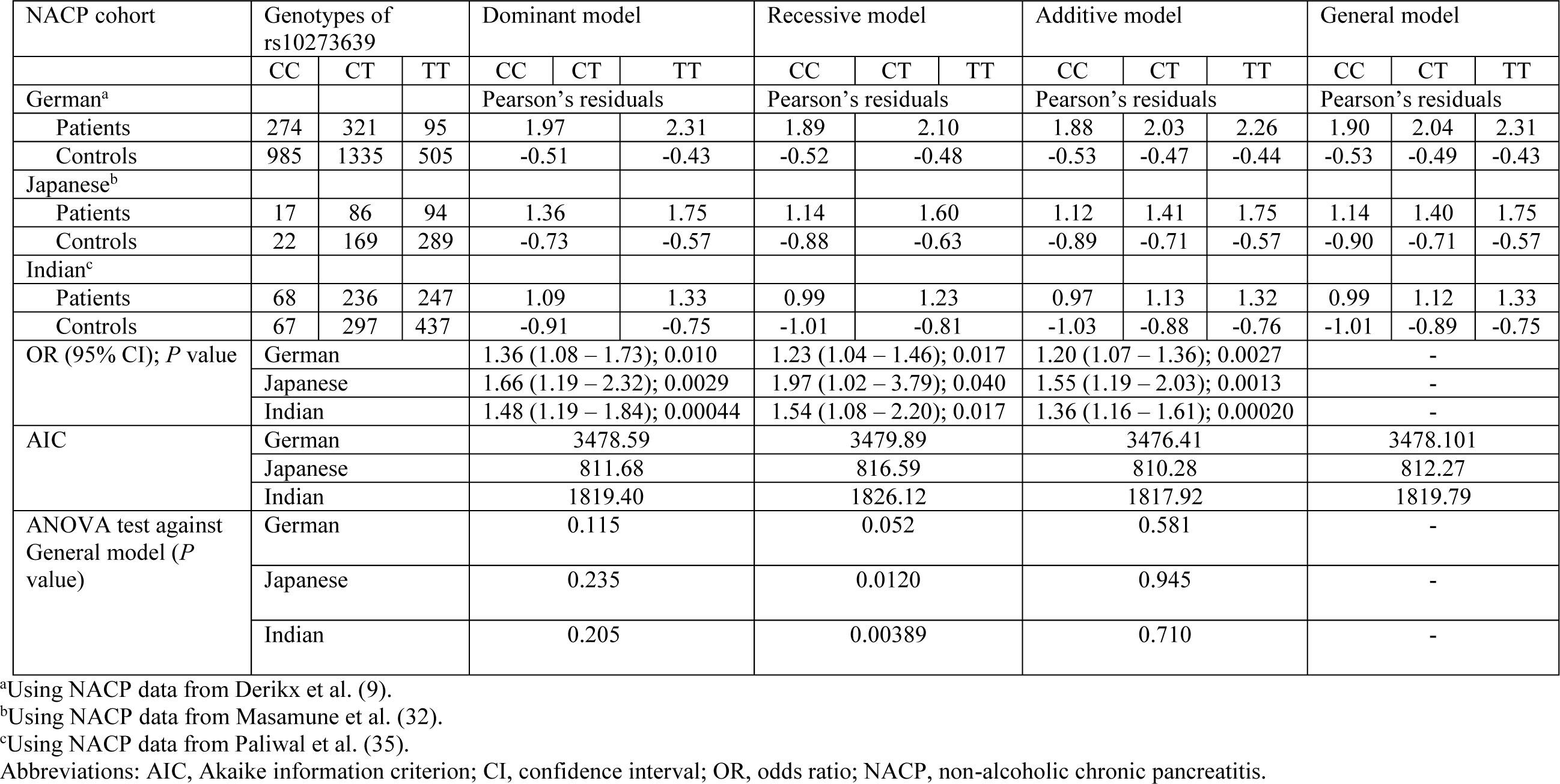
Test of the fit of different genetic models to the gene association data in three NACP cohorts

We also examined the three possible genetic models in the context of ACP studies. Of the five included ACP studies (Table 1), four used only healthy subjects as controls (6, 10, 32, 33). The Derikx study (9) additionally used German subjects with alcohol dependence (AD) and alcohol-associated liver cirrhosis (ALC) as controls. We therefore limited this analysis exclusively to the German data and tested the fit of the three genetic models to ACP association in the context of three control datasets. Again, the additive model provided the best fit to each dataset as evaluated by the AIC and by Anova tests against the general model. Pearson’s residuals did not indicate that the models fitted were unreasonable and in each dataset tested, a clearly significant correlation between genotypes of rs10273639 and the case/control status was observed as evidenced by the significant ORs (see Supplementary Table S1).

### A synergistic interaction is evident between the common *PRSS1-PRSS2* haplotype and alcohol consumption status

Using data from ACP and NACP patients from Germany, France, Netherlands, Japan and Hungary (Table 4), we first performed case-only re-analyses of each dataset to estimate the OR of the interaction between alcohol-consumption and copies of the *PRSS1/PRSS2* haplotype. Following the evaluation of different genetic models in NACP and ACP patients, we selected the best-performing additive genetic model for this re-analysis. Subsequently, we performed a meta-analysis of these new results in order to calculate a pooled estimate for the interaction term and to assess the heterogeneity of the studies. Application of the test for heterogeneity confirmed that the five studies were homogeneous (χ^2^ = 4.78, *P* = 0.31; I^2^ = 16.4%). The pooled OR (under a fixed effect model) of the interaction (or synergy index SIM) was 1.41 (95% CI 1.26–1.58; *P* = 2.80 × 10^−9^). The OR being significantly greater than one indicates that the effects between alcohol exposure and the haplotype are more than multiplicative (synergistic effect).

**Table 4.** Estimation of gene-environment interaction between the common *PRSS1-PRSS2* haplotype and alcohol consumption using a case-only analysis

| Population<sup>a</sup> | OR (95% CI)<sup>b</sup> | *P* value |
| --- | --- | --- |
| German | 1.43 (1.23 – 1.66) | $4.14 \times 10^{-6}$ |
| French | 1.54 (1.11 – 2.15) | 0.010 |
| Dutch | 1.84 (1.25 – 2.73) | 0.0022 |
| Japanese | 1.11 (0.83 – 1.49) | 0.48 |
| Hungarian | 1.34 (0.91 – 1.98) | 0.14 |
| Pooled (fixed effect model) | 1.41 (1.26 – 1.58) | $2.80 \times 10^{-9}$ |

<sup>a</sup>Original German, French and Dutch data used for analysis were from Derikx et al. (9). Original Japanese and Hungarian data used for analysis were from Masamune et al. (32) and Hegyi et al. (10), respectively.
<sup>b</sup>For the effect of the risk genotypes in the alcoholic chronic pancreatitis group against the non-alcoholic chronic pancreatitis group, equivalent to an estimate of the multiplicative-scale interaction term (or synergy index) between *PRSS1/PRSS2* and alcohol consumption.
Abbreviations: CI, confidence interval; OR, odds ratio.

To fully characterize the importance of the interaction, we fitted a multinomial logistic regression model to estimate, within a single framework, the risks associated with the *PRSS1-PRSS2* haplotype in both ACP and NACP cases (Table 5). For this analysis, control groups were required from each population. It should be noted that the purpose of the model is to compare the ACP and NACP against a common baseline and hence no assumption is made about alcohol consumption in the control groups. The multinomial model included co-variables indicating the country- and study-origin of the data, and we again applied the additive genetic model for the genetic effects. The OR for the *PRSS1/PRSS2* haplotype was 1.16 (95% CI 1.07–1.26) in the NACP patients and increased to 1.69 (95% CI 1.54–1.84) in the ACP patients. The difference between these two ORs is statistically significant, as evidenced by the non-overlapping confidence intervals, again signaling the presence of an interaction between the genetic-effect of the haplotype and alcohol consumption. This model allows for a succinct demonstration that alcohol consumption significantly increases the risk associated with the presence of copies of the haplotype on a multiplicative scale across the five populations.

**Table 5.** Test of gene-environment interaction between the common *PRSS1-PRSS2* haplotype and alcohol consumption using a multinomial logistic regression model

| Groups | Population<sup>a</sup> | Genotypes of the common haplotype<sup>b</sup> |  |  | OR (95% CI)<sup>c</sup> |
| --- | --- | --- | --- | --- | --- |
|  |  | CC | CT | TT |  |
| Controls | German | 985 | 1335 | 505 | – |
|  | French | 316 | 530 | 218 |  |
|  | Dutch | 146 | 222 | 66 |  |
|  | Japanese | 22 | 169 | 289 |  |
|  | Hungarian | 98 | 143 | 55 |  |
| NACP | German | 274 | 321 | 95 | 1.16 (1.07 – 1.26) |
|  | French | 130 | 189 | 96 |  |
|  | Dutch | 28 | 47 | 12 |  |
|  | Japanese | 17 | 86 | 94 |  |
|  | Hungarian | 44 | 42 | 17 |  |
| ACP | German | 433 | 358 | 73 | 1.69 (1.54 – 1.84) |
|  | French | 36 | 45 | 9 |  |
|  | Dutch | 115 | 102 | 15 |  |
|  | Japanese | 23 | 131 | 118 |  |
|  | Hungarian | 59 | 50 | 11 |  |

<sup>a</sup>Original German, French and Dutch data used for analysis were from Derikx et al. (9). Original Japanese and Hungarian data used for analysis were from Masamune et al. (32) and Hegyi et al. (10), respectively.
<sup>b</sup>Genotypes for the German, French, Dutch and Japanese cohorts refer to rs102373639 whilst those for the Hungarian cohort refer to rs6666.
<sup>c</sup>For the effect of the risk genotypes in the ACP or NACP group against the control group.

## DISCUSSION

We first performed an allele-based meta-analysis of the currently available studies of the association between the common *PRSS1-PRSS2* haplotype and ACP and NACP, demonstrating a significant association in both contexts. Possible causes underlying the previous contentious NACP findings included small sample size, ethnic and population differences in genetic predisposition to CP, and differences in patient selection. For example, using only German samples, Derikx and colleagues observed a trend towards higher genetic effect sizes in patient groups characterized by a later age of disease onset (9), a possible explanation for the lack of a significant association in the French cohort (see Table 2). More specifically, a significant association was found in the German patients whose age of disease onset was ≥ 20 years but not in those whose age of disease onset was 10-19 years or < 10 years; the age of disease onset was invariably < 20 years in all French patients. Additionally, it should be noted that *PRSS1-PRSS2* rs10273639 was included in a recent meta-analysis, in which ACP and NACP were analyzed together (38).

Having confirmed an association of the common *PRSS1-PRSS2* haplotype with NACP, we further explored the underlying biological basis of this association from two complementary standpoints. As mentioned earlier, the risk [C] allele of rs10273639C/T has been found to be positively associated with *PRSS1* mRNA expression in pancreatic tissue. More specifically, *PRSS1* expression levels were highest in rs10273639[C] homozygotes, intermediate in heterozygotes and lowest in rs10273639[T] homozygotes (6). However, the number of pancreatic tissue samples subjected to quantitative *PRSS1* mRNA expression analysis was limited. We therefore evaluated the effect of rs10273639 genotypes on pancreatic trypsinogen gene expression using the GTEx dataset and found the level of *PRSS2* mRNA expression to correlate directly with the dosage of the risk [C] allele of rs10273639. A role for *PRSS2* in CP is supported by four lines of evidence. Firstly, a loss-of-function *PRSS2* variant is known to be protective against CP (39). Secondly, CP-causing or predisposing trypsinogen copy number variants involve both *PRSS1* and *PRSS2* (5, 40). Thirdly, the *CTRB1-CTRB2* inversion increases CP risk by influencing protective PRSS2 degradation (30). Finally, transgenic expression of human *PRSS2* exacerbates pancreatitis in mice (41). A dosage effect of the risk allele on *PRSS1/PRSS2* expression in the pancreas implies that the additive gene model represents the best fit to explain the disease association data. Thus, the second approach we adopted was to test this hypothesis by a series of statistical analyses. We demonstrated that the additive genetic model provided the best fit in both the ACP and NACP contexts and the genetic effect of the rs10273639 CC genotype was approximately double that of the CT genotype. Taken together, these findings provide new insights into the pivotal role of increased trypsin in the etiology of CP as well as the genotype-phenotype relationship in ACP/NACP.

Alcohol is an established environmental risk factor for CP (3). However, only 3-5% of heavy drinkers develop the disease (42), suggesting the involvement of additional genetic or environmental factors in the causation of ACP. A higher OR of the common *PRSS1-PRSS2* risk allele associated with ACP than with NACP suggests a gene-environment interaction between the common haplotype and alcohol consumption in CP. By comparing allele distributions at rs10273639 between 447 ACP patients and 1,129 NACP patients, Whitcomb and colleagues (6) suggested the existence of an interaction between the *PRSS1-PRSS2* haplotype and alcohol although they did not attempt to quantify it. Herein, by fitting a similar but more sophisticated model and by combining inference from data of several studies, we confirm that there is indeed a synergistic interaction between the *PRSS1-PRSS2* haplotype and alcohol consumption status i.e. the risk associated with the presence of the *PRSS1-PRSS2* haplotype and exposure to alcohol was significantly greater than the product of the risks associated with each factor singly. Our results are in accordance with those of Whitcomb and colleagues but here, by first determining the correct genetic model (additive), and second by applying multinomial logistic regression, a more complete characterisation of the etiology of CP regarding the *PRSS1-PRSS2* haplotype and alcohol consumption was possible.

This study has several limitations. For example, we could not exclude publication bias in terms of some of the included studies. We did not perform funnel plots to assess this possibility following advice that such a test should be used only when there are at least 10 eligible studies; otherwise the power of the test is too low to distinguish a chance effect from genuine asymmetry (13, 14). Despite lacking detailed records of individual alcohol consumption levels, we have studied the role of alcohol by comparing ACP and NACP cohorts. We can be confident that these two groups will have experienced quite different levels of alcohol consumption, and hence the methods applied here are appropriate for detecting and describing a gene-environment interaction. More detailed data regarding the environmental exposure variable could enable more precision in the characterization of the interaction. Moreover, there may exist subtle differences between different cohorts in terms of ACP and NACP definitions. Furthermore, none of the included studies employed sex- and age-matched controls. However, we consider it unlikely that either of the abovementioned limitations would affect the main conclusions drawn owing to the mechanistic evidence underpinning the association data.

In conclusion, the results of our allele-based meta-analysis demonstrated significant association of the common *PRSS1-PRSS2* haplotype with both ACP and NACP. In addition, we have provided new information that sheds light on the pivotal role of uncontrolled trypsin in CP. Finally, we have refined the gene-environment interaction between the common *PRSS1-PRSS2* haplotype and alcohol consumption status in the etiology of CP. These findings help to improve our understanding of the complex etiology of CP and strengthen the notion that common risk factors (6, 30, 43) should be considered in risk assessment in the clinical setting. Acute pancreatitis patients carrying the common *PRSS1-PRSS2* haplotype should consider greatly reducing their alcohol consumption or abstaining altogether.

## Data Availability

All data relevant to the study are included in the article or uploaded as supplementary information.

## Author contributions

A.F.H. and E.G. designed and performed model testing and gene-environment interaction analysis and wrote the corresponding text. D.N.C. contributed to data interpretation and critically revised the manuscript with important intellectual input. E.M. assisted in performing the literature search and data extraction and contributed to the revision of the manuscript. C.F. contributed to the conception of the study, revised the paper, and provided overall supervision.

J.-M.C. conceived the study, performed the literature search, data extraction and meta-analysis, and wrote the paper. All authors have approved the final draft submitted.

## Acknowledgments

We are grateful to the authors who published studies that contributed data for our meta-and re-analyses. Appendix Figure 1 was obtained from the Genotype-Tissue Expression (GTEx) Portal; the GTEx Project was supported by the Common Fund of the Office of the Director of the National Institutes of Health, and by NCI, NHGRI, NHLBI, NIDA, NIMH and NINDS.

