## Supplementary Figure S1 for "Role of the common *PRSS1-PRSS2* haplotype in alcoholic and non-alcoholic chronic pancreatitis: meta- and re-analyses"

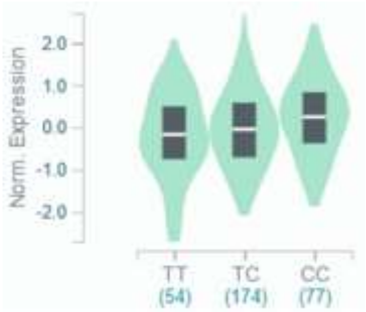


**Supplementary Figure S1**. Effect of the common *PRSS1-PRSS2* haplotype-tagging rs10273639C/T on *PRSS2* mRNA expression in the pancreas. C is the risk allele. Data were obtained from the Genotype-Tissue Expression (GTEx) Portal.
