## Supplementary Table S1 for "Role of the common *PRSS1-PRSS2* haplotype in alcoholic and non-alcoholic chronic pancreatitis: meta- and re-analyses"

**Supplementary Table S1.** Test of the fit of different genetic models to three German ACP association datasets*

| Association | Genotypes of rs10273639 | | | Dominant model | | | Recessive model | | | Additive model | | | General model | | |
|---|---|---|---|---|---|---|---|---|---|---|---|---|---|---|---|
| | CC | CT | TT | CC | CT | TT | CC | CT | TT | CC | CT | TT | CC | CT | TT |
| | | | | Pearson's residuals | | | Pearson's residuals | | | Pearson's residuals | | | Pearson's residuals | | |
| **Dataset 1** | | | | | | | | | | | | | | | |
| ACP patients | 433 | 358 | 73 | 1.71 | | 2.63 | 1.51 | 2.07 | | 1.50 | 1.96 | 2.56 | 1.51 | 1.93 | 2.63 |
| Healthy controls | 985 | 1335 | 505 | -0.58 | | -0.38 | -0.66 | -0.48 | | -0.67 | -0.51 | -0.39 | -0.66 | -0.52 | -0.38 |
| **Dataset 2** | | | | | | | | | | | | | | | |
| ACP patients | 433 | 358 | 73 | 0.97 | | 1.43 | 0.80 | 1.19 | | 0.81 | 1.10 | 1.50 | 0.81 | 1.13 | 1.43 |
| AD controls | 281 | 456 | 150 | -1.04 | | -0.70 | -1.24 | -0.84 | | -1.23 | -0.90 | -0.67 | -1.24 | -0.89 | -0.70 |
| **Dataset 3** | | | | | | | | | | | | | | | |
| ACP patients | 433 | 358 | 73 | 0.81 | | 1.29 | 0.72 | 0.98 | | 0.71 | 0.93 | 1.22 | 0.72 | 0.91 | 1.29 |
| ALC controls | 226 | 296 | 121 | -1.23 | | -0.78 | -1.38 | -1.02 | | -1.41 | -1.07 | -0.82 | -1.38 | -1.10 | -0.78 |
| OR (95% CI); *P* value | Dataset 1 | | | 2.36 (1.83 – 3.08); 7.5e-11 | | | 1.87 (1.61 – 2.19); 1.2e-15 | | | 1.70 (1.52 – 1.92); < 2e-16 | | | - | | |
| | Dataset 2 | | | 2.21 (1.64 – 2.98); 1.8e-7 | | | 2.17 (1.79 – 2.63); 6.4e-15 | | | 1.83 (1.59 – 2.13); 2.3e-16 | | | - | | |
| | Dataset 3 | | | 2.51 (1.85 – 3.44); 6.3e-9 | | | 1.85 (1.50 – 2.29); 9.1e-9 | | | 1.72 ( 1.48 – 2.01); 2.6e-12 | | | - | | |
| AIC | Dataset 1 | | | 3970.12 | | | 3956.1 | | | 3934.80 | | | 3936.334 | | |
| | Dataset 2 | | | 2402.35 | | | 2369.12 | | | 2360.65 | | | 2361.82 | | |
| | Dataset 3 | | | 2025.64 | | | 2026.78 | | | 2010.17 | | | 2011.12 | | |
| ANOVA against General Model (*P* value) | Dataset 1 | | | 2.20e-9 | | | 3.08e-6 | | | 0.496 | | | - | | |
| | Dataset 2 | | | 6.89e-11 | | | 0.0023 | | | 0.362 | | | - | | |
| | Dataset 3 | | | 4.8e-5 | | | 2.6e-5 | | | 0.307 | | | - | | |

*Genotype data from Derikx et al. (2015).

Abbreviations: ACP, alcoholic chronic pancreatitis; AD, alcohol dependence; AIC, Akaike information criterion; ALC, alcohol-associated liver cirrhosis; CI, confidence interval; OR, odds ratio.
